# An Interpretable Population Graph Network to Identify Rapid Progression of Alzheimer’s Disease Using UK Biobank

**DOI:** 10.1101/2024.03.27.24304966

**Authors:** Weimin Meng, Rohit Inampudi, Xiang Zhang, Jie Xu, Yu Huang, Mingyi Xie, Jiang Bian, Rui Yin

## Abstract

Alzheimer’s disease (AD) manifests with varying progression rates across individuals, necessitating the understanding of their intricate patterns of cognition decline that could contribute to effective strategies for risk monitoring. In this study, we propose an innovative interpretable population graph network framework for identifying rapid progressors of AD by utilizing patient information from electronic health-related records in the UK Biobank. To achieve this, we first created a patient similarity graph, in which each AD patient is represented as a node; and an edge is established by patient clinical characteristics distance. We used graph neural networks (GNNs) to predict rapid progressors of AD and created a GNN Explainer with SHAP analysis for interpretability. The proposed model demonstrates superior predictive performance over the existing benchmark approaches. We also revealed several clinical features significantly associated with the prediction, which can be used to aid in effective interventions for the progression of AD patients.

## Introduction

Alzheimer’s Disease (AD) is a complex, progressive neurodegenerative disorder that predominantly impacts older adults. It is estimated that approximately 6.7 million Americans 65 years or older have been diagnosed with AD^1^, leading to significant impacts on their quality of life and substantial burdens on the healthcare systems and caregivers. AD is a heterogeneous disease, with patients often exhibiting various rates of decline in cognitive status. For example, mild cognitive impairment (MCI) is a typical intermediate state between normal cognitive function and dementia due to AD^2^. Patients with MCI have a high probability of developing into AD with different progression timeframes, ranging from less than 1 year to as long as over 15 years^3,4^. Similarly, patients diagnosed with AD will also have a distinct survival time, with 1.5-3 years on average^5^. Therefore, identifying rapid progressors (RPs) with AD is critical for the prevention of the progression and clinical treatment. Previous studies have shown that multiple factors could accelerate the progression of AD, including age, genetics, and lifestyle^6^. Additionally, comorbidities such as hypertension, diabetes, and depression are also associated with RPs^7^, while social support may be protective in slow progression of AD^8^. A variety of assessments have been utilized to measure the cognitive decline, e.g., Mini-Mental State Examination (MMSE)^9^ and the 14-item cognitive subscale of the Alzheimer’s Disease Assessment Scale (ADAS-Cog14)^10^, which can be used to determine the progression speed of AD patients.

Previous research has employed various definitions to identify the rapid progression of AD. One of the most commonly used measurements is MMSE. Soto et al. demonstrated that the loss of equal to or greater than 4 points in MMSE score during the first six months of follow-up is a predictor of worse clinical course that can be used to define as a RP^11^. However, Schmidt et al. suggested that rapid progression of AD should be defined as MMSE score decrease of 6 points per year^12^. Another study showed that a mean MMSE score decrease of 5.5 points per year was defined as a rapid decline^13^. Some other methods have also been applied to define the rapid progression of AD, such as ADAS-Cog 13 and cerebrospinal fluid (CSF) biomarkers. For example, Ba et al. grouped rapid progression AD with 36.25 points decline in ADAS-Cog13^14^. Wallin et al. demonstrated patients who had low levels of CSF Aβ 1-42 (∼362±66 pg/mL), high levels of CSF total tau (1501±292 pg/mL), and high levels of CSF p-tau (139±39 pg/mL) decrease more quickly on cognition function and have higher mortality rate.

However, accurately identifying RPs of AD remains challenging, as there is no universal consensus on the definition of rapid AD progression and the measurement heavily relies on the availability and quality of the above-mentioned tests. The increasingly available and heterogeneous patient information from large-scale electronic health records (EHRs), as well as advancement in machine learning (ML) techniques^15–17^ makes it possible to reveal underlying patterns associated with RPs of AD. In this study, we hypothesized that the progression duration of patients from one diagnosis to another, e.g., from MCI to AD, is directly associated with the progression rate and can be used to infer the progression speed of AD. Here, we developed an interpretable population graph network to identify RPs of AD using diverse patient information (e.g., demographics, comorbidities and treatments) in EHRs from UK Biobank. The proposed graph-based structures can effectively integrate heterogeneous information from EHRs and capture associations among patients through edges. In particular, the construction of the population graph enables the modeling of clinical features to uncover the potential patterns of progression among the patients, facilitating better recognition of RPs. The overall workflow of our proposed framework is illustrated in **Figure 1**.

**Figure 1.**
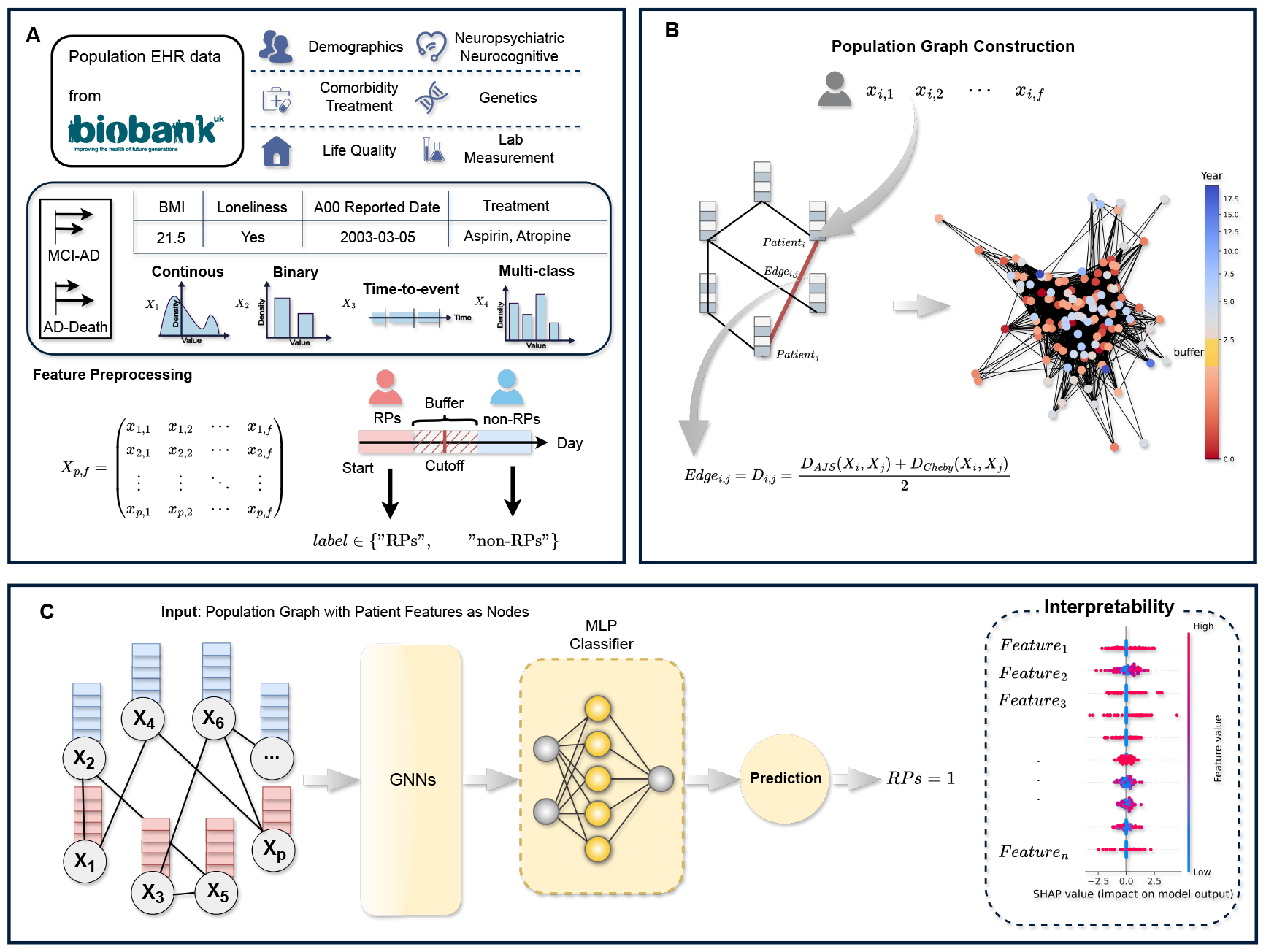
Illustration of the proposed framework. (**A**) The study participants from UK Biobank and patient feature selection. (**B**) Population graph construction. (**C**) Graph neural networks for identifying RPs of AD with interpretation.

## Materials and Methods

### Data source and participant selection

We used large collections of EHR data from UK Biobank^18^, a biomedical database and research resource from half a million UK participants, involving detailed patient information such as demographics, enrollment status, diagnoses, and lab results, etc. Here, we will have two different groups in the study to investigate AD progression, namely, from MCI to AD (MCI-AD) and from AD to death (AD-Death). We will use the following criteria to collect our cohorts: For patients in the MCI-AD group: (1) they have diagnosis dates of MCI and AD; (2) the diagnosis date of MCI is earlier than the diagnosis date of AD. For patients in the AD-Death group: (1) they have a diagnosis date of AD and death time; (2) the diagnosis date of AD is earlier than the death date. The diagnosis of MCI and AD was identified with ICD codes, that is, ICD-10 codes F06 and G31 for MCI, and ICD-10 codes F00 and G30 for AD. We ended up with 177 and 979 cases for MCI-AD and AD-Death groups, respectively.

### Problem formulation

To identify RPs of AD using EHR data, in this paper, we define the progression speed of AD based on the duration of the patient diagnosis from one status to another. Previous work has shown that, on average, 548-730 days (1.5-2 years) is a relatively reasonable observation window for the cognitive decline conversion from MCI to AD^19,20^. Similarly, the observation window for patient survival time after being diagnosed with AD is around 548-1095 days (1.5-3 years)^5,21^.

Considering the progression time distribution of the study cohort at two different groups, we selected 660 days and 889 days as cut-offs for MCI-AD and AD-Death groups, respectively, which were the mean values of progression duration in the two study cohorts. However, it is challenging to determine if the AD patient is in rapid progression only using a single cut-off day as a metric. We introduced “buffer” to expand the duration based on our selected cut-off day to define the fast progression of AD patients. For example, in the MCI-AD group, we select the cut-off day as 660 and we introduce the ‘buffer’ ratio, a weight ranging from 0 to 1 to calculate ‘buffer’ day. Here, we set = 0.2, The ‘buffer’ day is equal to 0.2 * cut-off day, which is 132. As a result, we define the patient with the time from MCI to AD shorter than 528 (cut-off day – “buffer” day) as RP, likewise, the patient with the time greater than 792 days (cut-off day + “buffer” day) is labeled as non-RP. To reduce the bias in the analysis, we will exclude the patients whose transition time of MCI-AD is within [528, 792] and use the remaining cases for modeling to identify RP or non-RP patients of AD. To demonstrate the robustness of this formulation, we also experimented with different values of the “buffer” ratio to examine and compare the prediction results.

### Feature preprocess and selection

The clinical information of selected patients in two cohorts was extracted from UK Biobank electronic health-related records, which consists of thousands of different features. We categorize these features into 4 types, i.e., continuous features (e.g., body mass index), binary features (e.g., sex), time-to-event features (e.g., date of attending assessment center) and multi-class features (e.g., medication code). We first transformed and normalized these features with different strategies based on categories to ensure they can be processed by the classifiers. We utilized Min-Max normalization to convert continuous features with a distribution between 0 and 1. For binary and multi-class categorical features, we applied one-hot method to encode these variables. The time-to-event features are converted into numerical values using the information at different time points. We then removed the features with missingness rate over 30% and applied four different feature selection methods, including variance threshold, Pearson Correlation, Chi^2^ test and ANOVA test, to further filter out the remaining features that were less relevant to our predictive targets by the following rules (1) Variance threshold < 0.1; (2) Pearson Correlation between features and targets < 0.01; (3) P-value >= 0.05 by Chi^2^ test; and (4) P-value >= 0.05 by ANOVA test. Ultimately, we obtained 169 and 177 features in MCI-AD and AD-Death groups, respectively, for the construction of the model. The remaining features can be divided into 6 different categories involving demographics, neuropsychiatric and neurocognitive, comorbidity and treatment, lab measurements, genetics and life quality. The details of these features can be found in Supplementary Material S1. Lastly, we employed the k-nearest (k = 3) neighbor to impute the missing values on selected features.

### Population graph construction

Graph-based methods have been applied to neurodegenerative disease studies, including AD^22^, as graphs can inherently manage multimodal information by integrating the patients’ diverse medical information as node features through a similarity metric. The structure of the graph is extremely crucial for combining various information and downstream tasks. It has been shown that a simple multi-layer perceptron (MLP) can outperform a complex graph neural network with a poor structure^23^. Therefore, to construct a robust graph, we introduced population graphs to represent two study cohorts of AD patients involving their clinical information. We use the MCI-AD group as an example to describe the construction of the population graph. Assume we have the patient dataset 𝔻 = {(*x*_*i*_, *y*_*i*_) | *x*_*i*_ ∈ *X, y*_*i*_ ∈ *Y*}, where *x*_*i,j*_ ∈ ℝ^*p*×*f*^ is the input matrix and each patient can be represented as *X*_*i*_ = *x*_*i*,1,_ *x*_*i*,2, ⋯,_ *x*_*i,f*_ and *f* is the number of features. A population graph has the structure as 𝒢 = {𝒱, ℰ}, where 𝒱 is the set of nodes, one per patient, and ℰ is a set of edges connecting the paired nodes. We use distance *D* to judge whether there is an edge between two patients and the value of *D* is used as the edge weight *E*_*i,j*_. If two patients’ distance is under a threshold *θ* then we add an edge (*e*_*i*_, *e*_*j*_) with the weight *E*_*i,j*_. We looped the distance calculation and edge-adding processes and accumulated the *θ* by step *η* until we had a connected population graph 𝒢.

In terms of the calculation of *D*, we leveraged the average of two types of features’ distance between patients. From the process in the feature selection, we know that the binary and multi-class features belong to categorical variables, while the continuous and transformed time-to-event features are numerical variables. For a rigorous computation of the distance *D* and edge weight *E*_*i,j*_, we propose a novel approach utilizing the weighted sum of Average Jaccard Similarity Distance *D*_*AJS*_ and Chebyshev Distance *D*_*Cheby*_ to compute the distance between patients. Here, the *D*_*AJS*_ can effectively measure the distance between categorical variables^24^, and the *D*_*Cheby*_ can swiftly calculate the maximum distance between numerical variables^25^. We used their average value to calculate a distance for paired patients, the formulas are presented as follows:

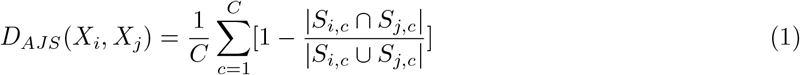

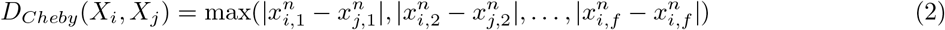

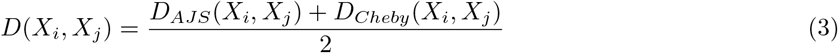

where *C* represents the total number of categorical variables, *n* is the numerical variables. *S*_*i,c*_ and *S*_*j,c*_ denote the sets of categorical variables and 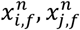 represent the numerical variables for patients *X*_*i*_ and *X*_*j*_ respectively. A detailed description of constructing the population graph steps is as follows:

#### Algorithm Population Graph Construction

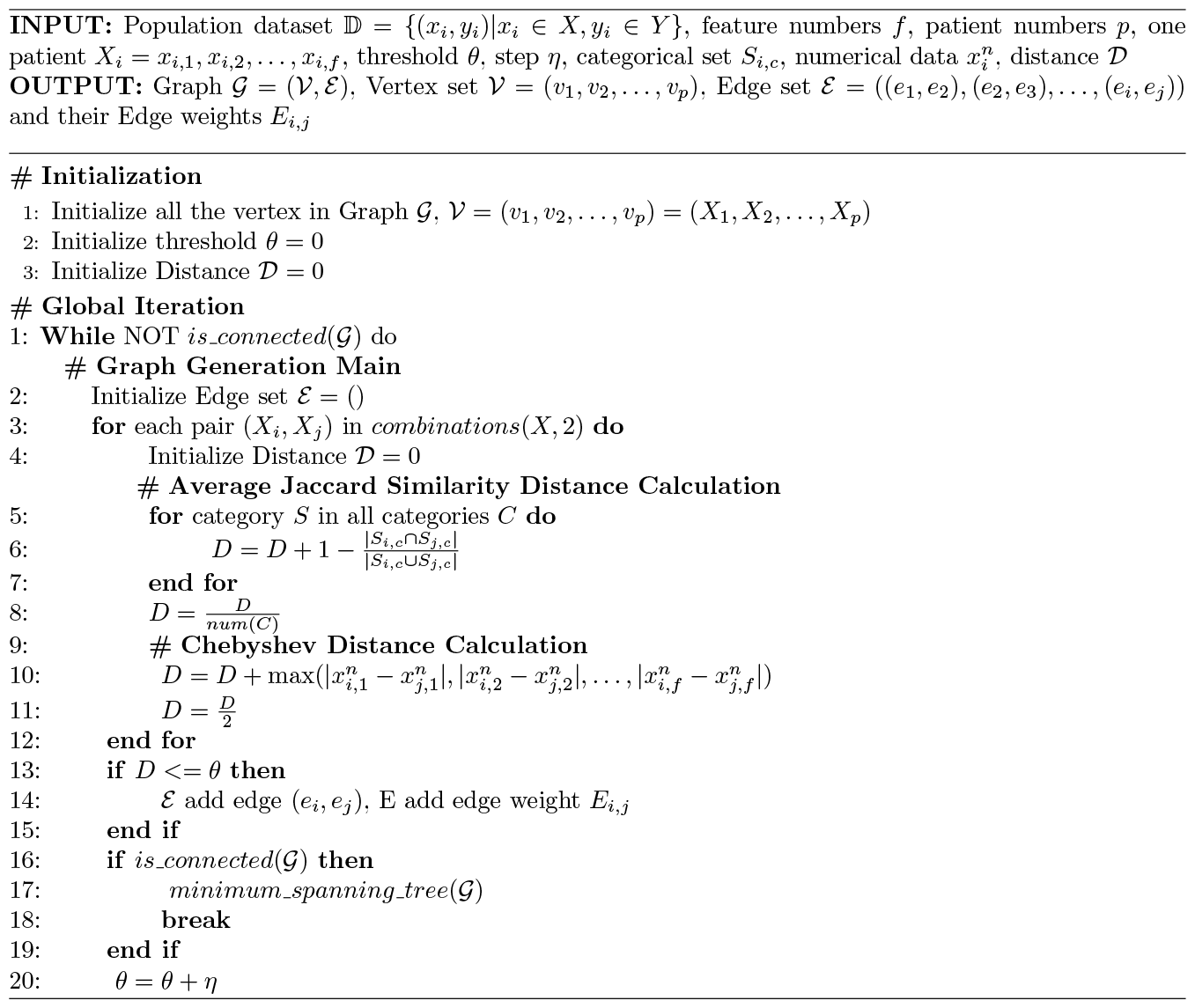

### Graph neural networks

To further identify the RPs of AD patients denoted as nodes in the population graph, we introduced graph neural networks (GNNs) capable of learning representations from graph-structured data. The GNNs leverage the graph structures to iteratively propagate and aggregate node features, mathematically formulated as:

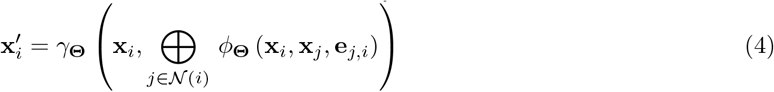

In this equation, 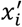 denotes the new representation of node *i*, which is a patient in this case, *𝛾*_*Θ*_ is a learnable aggregation function, ⊕ represents the aggregation operation, 𝒩_(*i*)_ is the set of neighbors of node *i, ϕ*_*Θ*_ is a learnable transformation function, and *e*_*j,i*_ represents the feature of the edge from node *j* to node *i*. Each node was initialized with a feature vector representing clinical information (e.g., demographics, lab results) of a patient in the population graph. The GNNs were leveraged to process and aggregate the feature information from neighbor nodes. Through the integration of multiple GNN layers, we can implement inter-node message passing and generate the final graph encoding vector by calculating the average value across aggregated embeddings. Finally, we fed the graph encoding vector to a fully connected layer with a sigmoid activation function for the prediction. Here, we employed three types of GNN architectures, namely, graph sample and aggregate network (SAGE), graph convolutional network (GCN), and graph attention network (GAT), to identify RPs in AD in our two study cohorts.

### Model interpretation

Understanding the mechanism of how the classifier makes a confident prediction is equally vital as predictive performance, particularly in biomedical fields. Here, we aim to investigate the feature importance of GNNs for RPs prediction of AD in our two different study cohorts. We implemented a saliency analysis using a GNN Explainer^26^ that combines the interpretability of graph neural networks with the explainability provided by SHAP^27^ (SHapley Additive exPlanations) values. We trained a GNN model to predict RP of AD and created an explainer model alongside the GNN. We then used the GNN Explainer model to generate SHAP values for each node and edge in the input graph involving perturbing the graph and observing the effect on the model’s predictions to calculate the SHAP values. The SHAP values were aggregated for each node and edge to determine the overall impact on the model’s predictions. Based on the results, we can obtain how much each attribute contributes, either positively or negatively, to the target label. By applying SHAP values, we could obtain masks for each node in the population graph, containing information about the importance of both the node and its features. Since previous studies mainly relied on scores such as MMSE for classifying RPs, while our study included more categories of features, it is essential to figure out how these features contribute to the decision-making process for RP identification in AD.

### Experimental setup Implementation and evaluation

The dataset of each group (MCI-AD, AD-Death) was randomly divided into three sets: training (70%), validation (10%), and testing (20%). The RPs of AD were labeled as “1” and non-RPs as “0”, representing positive and negative samples. Our GNN-based models were optimized across 100 training epochs with a batch size of 16, a learning rate of 0.001, and a dropout strategy with a rate of 0.2. We trained our model with 5-fold cross-validation on the training set, fine-tuned and evaluated it on the validation and testing set separately. To measure the models’ prediction performance, we employed five distinct evaluation metrics, including accuracy, precision, recall, F1-score, and the area under the receiver operating characteristic curve (AUROC). In addition, we also investigated whether the different values of ‘buffer’ day could influence our models’ prediction performance.

### Benchmark approaches

To evaluate the performance of the proposed framework for identifying RPs of AD in each group, we conducted comparative experiments using several benchmark approaches involving classic ML models, e.g., logistic regression (LR), random forest (RF), and XGBoost, and deep neural networks (DNNs), e.g., multilayer perceptron (MLP), bidirectional long short-term memory (BiLSTM) and residual neural network (ResNet). The parameters and hyperparameters of these benchmark models can be found in Supplementary Material S2. The datasets used to train and test these models are identical to our GNN-based structure to ensure a fair comparison process.

## Results

### Comparative performance between our models and benchmark approaches

**Table 1** shows the comparison performance for predicting RPs of AD patients through our proposed and benchmark approaches on two different groups. For the MCI-AD group, we utilized a cutoff of 660 days, with the optimal being 0.4, resulting in a total ‘buffer’ days of 347 days. Therefore, the patients with progression duration from MCI to AD in 313 days are classified as RPs (78 cases), likewise, the patients with duration over 1,007 days are labeled as non-RPs (56 cases). From the results, we could observe that DNNs outperformed traditional ML classifiers for the prediction generally. The BiLSTM showed the best performance in terms of recall (1.000) and AUROC (0.976), though MLP also achieved decent and comparative results on accuracy (0.929), recall (1.000), and F1-score (0.933). When we compared our graph models with others, we found that SAGE is the best architecture among GNNs and displays comparable performance with 0.929 in accuracy, 1.000 in precision and 0.976 in AUROC. Due to the relatively small cohort we trained, the simple aggregation approach in SAGE, which weights and sums neighboring nodes, ensures sufficient feature extraction while reducing the impact of overfitting. We also noticed that GAT achieved the highest Recall (1.000), but its performance in other metrics was relatively poor. This might be attributed to GAT’s tendency to aggregate neighboring nodes with the highest contribution to the positive samples using attention mechanisms, which could lead to a decrease in the performance of other metrics.

**Table 1.** Performance comparison of predicting RPs of AD between our proposed framework and baseline classifiers in two different groups.

| Group | Model | Accuracy | Precision | Recall | F1-score | AUROC |
| --- | --- | --- | --- | --- | --- | --- |
| MCI-AD | LR | 0.815 | 0.875 | 0.824 | 0.848 | 0.812 |
|  | RF | 0.593 | 0.650 | 0.765 | 0.703 | 0.532 |
|  | XGBoost | 0.741 | 0.778 | 0.824 | 0.800 | 0.712 |
|  | MLP | **0.929** | 0.909 | **1.000** | **0.952** | 0.950 |
|  | BiLSTM | 0.923 | 0.875 | **1.000** | 0.933 | **0.976** |
|  | ResNet | 0.714 | 0.636 | **1.000** | 0.778 | 0.714 |
|  | SAGE | **0.929** | **1.000** | 0.857 | 0.923 | **0.976** |
|  | GCN | 0.857 | **1.000** | 0.714 | 0.833 | 0.918 |
|  | GAT | 0.786 | 0.700 | **1.000** | 0.824 | 0.959 |
| AD-Death | LR | 0.725 | 0.681 | 0.671 | 0.676 | 0.718 |
|  | RF | 0.614 | 0.613 | 0.606 | 0.609 | 0.639 |
|  | XGBoost | 0.643 | 0.611 | 0.452 | 0.520 | 0.619 |
|  | MLP | 0.686 | 0.700 | 0.538 | 0.609 | 0.715 |
|  | BiLSTM | 0.640 | 0.625 | 0.513 | 0.563 | 0.704 |
|  | ResNet | 0.597 | 0.593 | 0.504 | 0.549 | 0.628 |
|  | SAGE | 0.826 | 0.853 | 0.744 | 0.795 | 0.911 |
|  | GCN | **0.930** | **0.971** | 0.872 | **0.918** | **0.983** |
|  | GAT | 0.895 | 0.857 | **0.923** | 0.889 | 0.934 |

For the AD-Death group, we selected 889 days as the cutoff day and the best is 0.3 with ‘buffer’ of 287 days. Similarly, we collected 348 cases labeled as RPs (duration within 602 days) and 505 cases of non-RPs (duration over 1,176 days). The results of the AD-Death group indicate that our proposed population graph networks outperformed all other baseline approaches, showing an obvious superiority for the prediction. Among these, the population graph network with GCN architecture exhibited the best accuracy (0.930), precision (0.971), F1-score (0.918), and AUROC (0.983), while it obtained the highest recall (0.923) with GAT. Although the prediction performance by the SAGE model in the AD-Death group is not as good as in the MCI-AD group, it still exceeded 19.3% in AUROC compared with the best benchmark method, demonstrating the effectiveness of population graph structure with GNN models in identifying RPs through extracted the features. Moreover, we found that almost all the models presented a better performance for predicting RPs of AD in MCI-AD group than in AD-death group, despite the former group having a smaller cohort size. This is probably due to both cohorts only involving a relatively small sample size, while the patients in the AD-Death group would have a more diverse and sporadic distribution of features, which makes it more difficult to differentiate rapid and slow progression AD patients. Nevertheless, our proposed framework proved the effectiveness of identifying RPs of AD in both groups with superior performance over the existing benchmark methods.

### Ablation study

To investigate the impact of ‘buffer’ values on the models’ performance, we assess various ‘buffer’ ratios to see how they could affect the predictions of our models. Considering that both excessive small (days < 50) or large (days > 800) ‘buffer’ days might adversely affect the availability and quality of the study cohort in this work. We only selected 4 different values of the ‘buffer’ ratio in the experiments, that is, 0.2, 0.3, 0.4 and 0.5. We chose SAGE and GCN as our models in MCI-AD and AD-Death groups, respectively, which demonstrate the best performance on RP prediction, to examine the results with different ‘buffer’ days. The parameters and hyperparameters remained the same as in **Table1**. As shown in **Table 2**, in the MCI-AD group, when selecting, the model exhibited better performance over other values of on average, with the best accuracy (0.929), precision (1.000), F1-score (0.923), and AUROC (0.976). In the AD-Death group, we would obtain the same results when or, showing the highest accuracy (0.938), precision (1.000) and F1-score (0.931), which may suggest that the distribution of selected patients in RP and non-RP classes are the same in these cases. As a comparison, the model will have the best recall (0.872) and AUROC (0.983) values when. We also studied the impact of ‘buffer’ with other models and the results can be found in Supplementary Material S3. From all these results, we can conclude that the selection of ‘buffer’ ratio could influence the models’ prediction to some extent, but there are no significant changes on the prediction performance.

**Table 2.**
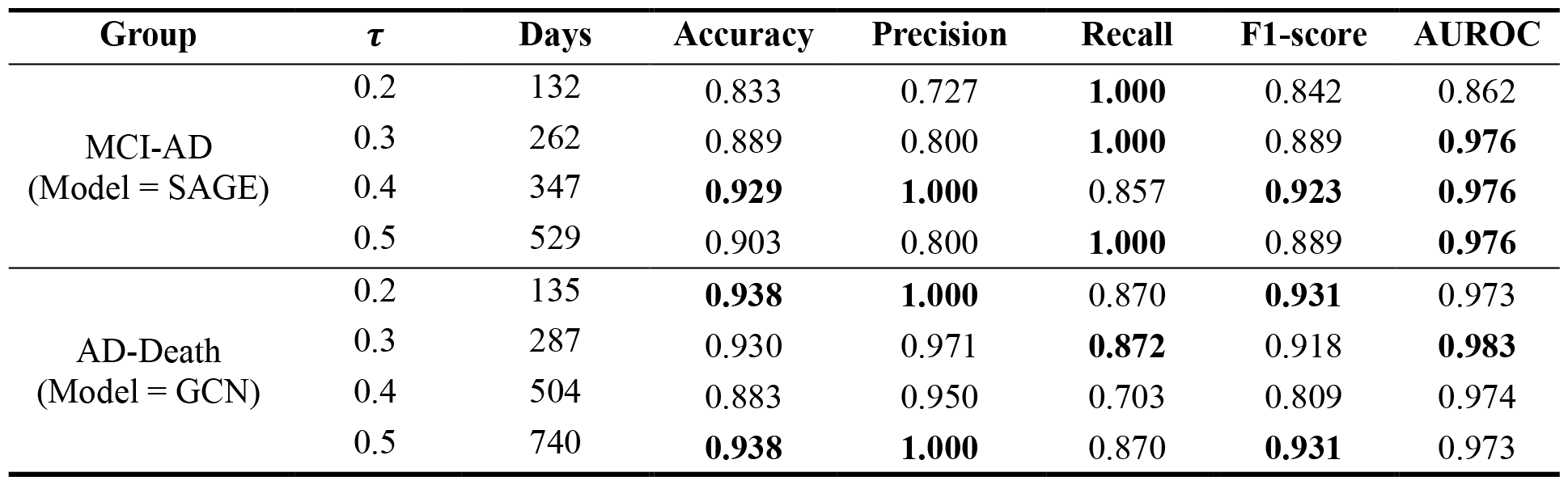
Performance comparison with different ‘buffer’ ratio for RPs prediction of AD in two study groups.

| Group | $\tau$ | Days | Accuracy | Precision | Recall | F1-score | AUROC |
| --- | --- | --- | --- | --- | --- | --- | --- |
| MCI-AD<br>(Model = SAGE) | 0.2 | 132 | 0.833 | 0.727 | **1.000** | 0.842 | 0.862 |
|  | 0.3 | 262 | 0.889 | 0.800 | **1.000** | 0.889 | **0.976** |
|  | 0.4 | 347 | **0.929** | **1.000** | 0.857 | **0.923** | **0.976** |
|  | 0.5 | 529 | 0.903 | 0.800 | **1.000** | 0.889 | **0.976** |
| AD-Death<br>(Model = GCN) | 0.2 | 135 | **0.938** | **1.000** | 0.870 | **0.931** | 0.973 |
|  | 0.3 | 287 | 0.930 | 0.971 | **0.872** | 0.918 | **0.983** |
|  | 0.4 | 504 | 0.883 | 0.950 | 0.703 | 0.809 | 0.974 |
|  | 0.5 | 740 | **0.938** | **1.000** | 0.870 | **0.931** | 0.973 |

### Model interpretation

We visualized the SHAP values of the top 20 important features for our models predicting RPs of AD in each study group. In the SHAP values for the MCI-AD group (**Figure 2**), we can identify several features with potential clinical significance for the progression of AD. For example, “Chest pain or discomfort_No”, which ranks the uppermost among all the features for the prediction, suggests a potential link between cardiovascular health status and the rate of AD progression. “Mood swings_Yes” is revealed as another significant feature, with a positive SHAP value indicating that mood fluctuation could be associated with rapid progression of AD. Also, “Interpolated Year when operation took place” highlights the importance of surgery timing in the model’s predictions. Similarly, two other features, i.e., “Urate” and “Haematocrit percentage” suggest their impact on AD progression, which is consistent with previous evidence that uric acid levels and hematocrit percentage are risk factors to cognitive impairment^28^. Similarly, we found that “Alcohol intake frequency_Once or twice a week” also affects model’s predictions, which may implicate the association between alcohol intake and AD progression. The feature “Operative procedures - OPCS4_U05.1 Computed tomography of head” emerges as another influential procedure-relevant feature, likely due to the direct information provided by head CT scans about brain health status. Regarding “Salt added to food_Sometimes,” the distribution of SHAP values suggests varying effects on AD progression, reflecting potential relationships between dietary habits and AD progression rate.

**Figure 2.**
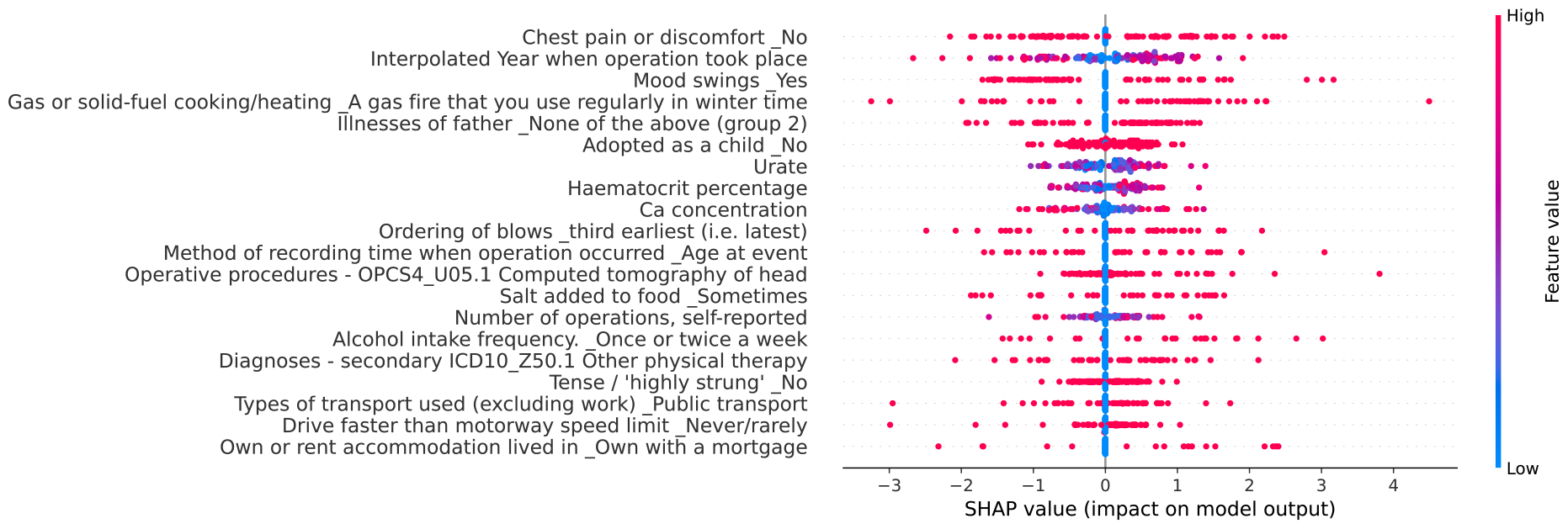
Summary plot between SHAP values and their impact on the RP prediction in MCI-AD group.

Correspondingly, **Figure 3** displays the SHAP values of top 20 features for predicting RPs of AD with our model in AD-Death group. From this figure, we can see that the most critical features are “Sleeplessness/insomnia_Sometimes”, “Sensitivity/hurt feelings_No” and “Diagnoses-secondary ICD10_E86 Volume depletion”. “Sensitivity/hurt feelings_No” is negatively associated with the model for identifying rapid AD progression, while the other two features are positive related. Meanwhile, we observed that “Genetic sex_Female” and “Sex_Male” are both important features. However, the SHAP values of these two features suggest that “Genetic sex_Female” has an opposite contribution to that of feature “Sex_Male”, implying that males are positively associated with the model identifying a rapid progression in the AD-Death group, whereas females are not. Additionally, “Salt added to food_Never/rarely” shows a significant negative SHAP value, indicating that a low-salt diet may be associated with a slower disease progression. Medical procedures related to monitoring, such as “Operative procedures - main OPCS4_U05.1 Computed tomography of head” and “Operative procedures - OPCS4_Y98.1 Radiology of one body area (or < 20 minutes),” remain correlated with disease progression rate. Moreover, the features “Diagnoses - secondary ICD10 R29.6 Tendency to fall, not elsewhere classified” and “Diagnoses - secondary ICD10 R26.8 Other and unspecified abnormalities of gait and mobility” suggest that difficulties in walking and a tendency to fall may serve as indicators of accelerated AD progression. The SHAP value of “Underlying (primary) cause of death: ICD10_F03 Unspecified dementia” positively impacts the model, indicating that other unspecified dementia-related factors play an important role in disease progression. Interestingly, “HDL cholesterol” is identified by the model as a feature with a negative impact, suggesting that high-density lipoprotein cholesterol levels may be associated with the progression rate in AD.

**Figure 3.**
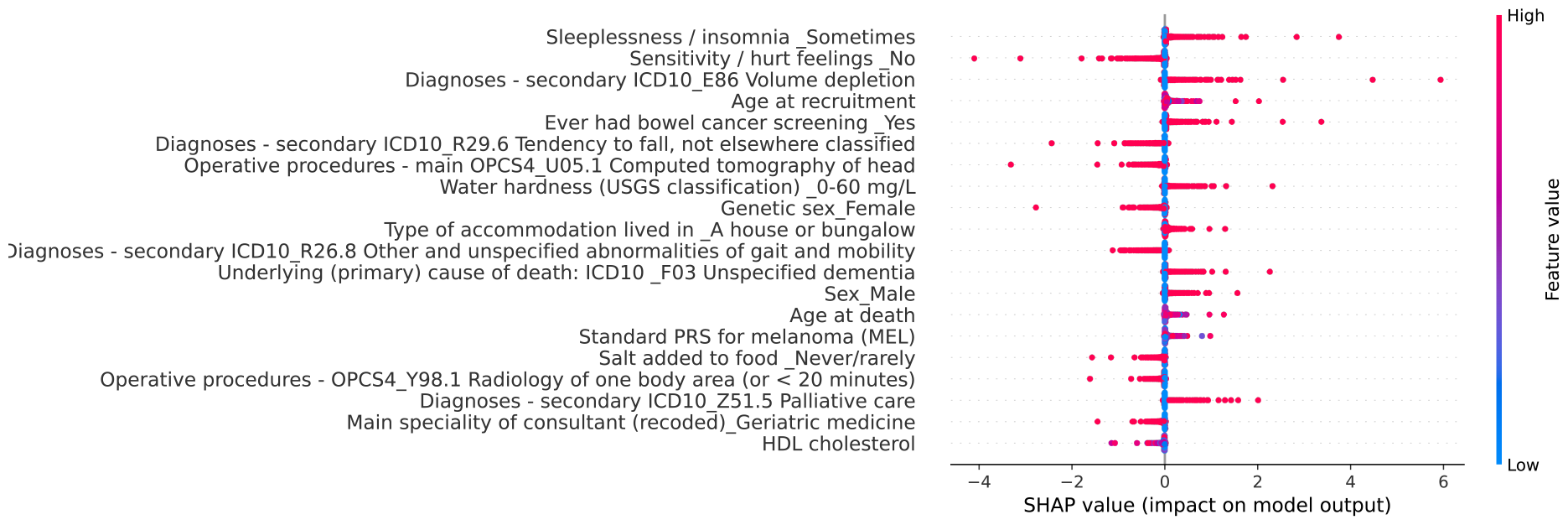
Summary plot between SHAP values and their impact on the RP Prediction in AD-Death group.

## Discussion and Conclusion

In this paper, we proposed an interpretable graph-based framework for predicting the rapid progression of AD, utilizing diverse patient information in EHRs from UK Biobank. We first defined the RPs and non-RPs of AD based on the duration of two different diagnoses of the patient, e.g., MCI to AD. We introduced ‘buffer’ (details in Materials and Methods) to reduce the bias for the definition of RP cases. The clinical information and features of selected patients were extracted and further processed from EHRs using several feature selection techniques to obtain the most relevant ones. We then employed a distance-based algorithm to construct population graphs on selected cohorts with extracted features of patients. Three different GNN structures were utilized to identify RP in AD patients. This framework was trained and evaluated on two different study cohorts we created, i.e., MCI-AD and AD-Death. In both cohorts, we successfully identified RP of AD using our proposed graph-based models. Compared with benchmark methods, our models achieved a better performance for RP prediction, especially in the AD-Death group. For instance, our proposed population graph model with GCN structure shows the best accuracy (0.930), precision (0.971), F1-score (0.918) and AUROC (0.983) that are 28.3%, 42.3%, 35.8% and 36.9% greater than the best benchmarks. Though the other two graph-based models are not as good as GCN, they outperformed all benchmark models in all evaluation metrics. The results in the MCI-AD group are a little different from the AD-Death group, that is, SAGE shows better prediction results than the other two graph models. However, in benchmark models, MLP and BiLSTM also display comparable performance with SAGE model that exceeds others. We also investigated how the ‘buffer’ would affect the prediction performance of our models. The experimental results turn out that we will gain the best prediction results when the ‘buffer’ ratio is 0.4 in MCI-AD and AD-Death. Despite that, the selection of ‘buffer’ to a reasonable extent does not have a very significant impact on the model performance.

Furthermore, we employed SHAP to explain our GNN models, providing interpretable analysis for our models’ predictions. We found that the important features discovered in the two cohort groups are quite distinct. This is primarily because the features extracted to build the model are based on the characteristics of the patients in each group, which are mostly non-overlapped. Therefore, the features identified as important for the prediction of RPs in AD vary a lot from these two groups. According to the results (**Figure 2** and **Figure 3**), “Chest pain or discomfort_No”, “Interpolated Year when operation took place” and “Mood swings_Yes” are the top 3 important features in MCI-AD group, whereas “Sleeplessness/insomnia_Sometimes”, “Sensitivity/hurt feelings_No” and “Diagnoses-secondary ICD10_E86 Volume depletion” are more important in AD-Death group for the prediction of RPs in AD. It is of note that among these, “Sensitivity/hurt feelings_No” is negatively associated while others are positively related. Some additional biomarkers identified by our models include “Urate” and “Haematocrit percentage”, “Alcohol intake frequency_Once or twice a week” and “Salt added to food_Sometimes,” in the MCI-AD group and “Genetic sex_Female”, “Sex_Male”, “Underlying (primary) cause of death: ICD10_F03 Unspecified dementia” in AD-Death group. Our models’ interpretable results are aligned with some previous studies, for instance, age has consistently been identified as an essential factor contributing to cognitive decline in Alzheimer’s Disease (AD)^6^. The gender differences results have also been noted in AD-related mortality studies, with females at higher risk but males exhibiting a faster rate of decline^29^. In terms of neurophysiology, Perneczky et al. observed that symptoms such as insomnia, depression, and hallucinations are associated with and accelerate cognitive decline during the progression of AD^2^. As for comorbidities, higher urate levels have been identified as protective factors against AD progression^28^. Soto et al. found a correlation between cardiovascular diseases in elderly individuals and AD, suggesting that repeated cardiovascular examinations might be a result of rapid AD progression^11^. It has also been revealed that higher salt intake could lead to declining performance in high-risk populations^30^. Overall, we contribute to the prediction of AD progression rates by proposing new methods and insights while providing interpretable and accurate predictions that could aid in the monitoring and prevention of AD progression.

There are several limitations of this study. Firstly, the study cohort in both groups is relatively small which might affect the models’ prediction ability. Also, all the patients were obtained from UK Biobank, which could limit the generalizability of our model when applied to AD patients with distinct characteristics (e.g., demographics) from other sites for rapid progression identification. Secondly, this study used progression time between records of different diagnoses to define whether it is a rapid or slow progression based on the duration. It may not be a gold standard to measure the cognitive decline to determine the progression rate, as the date of diagnosis in records for MCI or AD will not completely reflect the true onset time of the diseases. This could cause some bias when labeling the patients in terms of their progression rate for modeling. Additionally, age is a crucial factor for the progression rate of AD patients that could influence the definition of rapid progression in this work. Thirdly, this work only used EHRs for identifying RPs of AD. Though there is plenty of patient-related information, the use of EHRs overlooks other critical information that impact the diagnostics, e.g., cognitive assessments and clinician preferences, which could potentially improve the reliability of the predictions. Moreover, the EHRs may contain errors or glitches that could influence the integrity and accuracy of the collected information from the patients. The limitations highlight the importance of addressing these issues to improve the performance and generalizability of identifying the rapid progression of AD.

The future work of this study includes increasing the cohort size and improving the quality of the patient information to construct the prediction model that can more accurately identify RPs in AD and relevant critical features responsible for the progression rate. We will also incorporate several cognitive assessments, e.g., MMSE, in combination with the duration between diagnoses of patients that could provide a ground truth to determine the progression rate of patients with AD. This would reduce the bias or errors that a patient is labeled incorrectly before the construction of the model. Furthermore, to evaluate the generalizability of the predictive models for identifying RPs of AD, we will collect patient samples from multiple different sites for testing. We will also create a federated learning^31^, a machine learning approach that protects privacy by preventing the local accumulation of raw clinical data across various institutions. This would allow researchers to combine the data from different institutional sites while preserving patients’ privacy when predicting rapid progression for AD patients. In sum, our study aims to uncover the AD patients who are likely to experience rapid progression that can facilitate the early detection and prevention of the disease for patients.

## Data Availability

All data produced in the present study are available with access fee upon reasonable request to https://www.ukbiobank.ac.uk/. The data can be accessed via the UKB research analysis platform (RAP): https://ukbiobank.dnanexus.com/landing.

https://github.com/UF-HOBI-Yin-Lab/ADGraph/

## Supplementary Materials

The codes and supplementary materials are publicly available at: https://github.com/UF-HOBI-Yin-Lab/ADGraph/.

## Acknowledgements

This study was partially supported by grants from Centers for Disease Control and Prevention (1U18DP006512), National Institute of Environmental Health Sciences (R21ES032762) and the NIH National Center for Advancing Translational Sciences (UL1TR001427). This research has used the UK Biobank data (Application Number 95030).

